# GSTP1 positive prostatic adenocarcinomas are more common in Black than White men in the United States

**DOI:** 10.1101/2020.08.28.20183954

**Authors:** Igor Vidal, Qizhi Zheng, Jessica L. Hicks, Jiayu Chen, Elizabeth A. Platz, Bruce J. Trock, Ibrahim Kulac, Javier Baena-Del Valle, Karen S. Sfanos, Sarah Ernst, Tracy Jones, Stephanie Glavaris, William G. Nelson, Srinivasan Yegnasubramanian, Angelo M. De Marzo

## Abstract

GSTP1 is a member of the Glutathione-S-transferase (GSTS) family silenced by CpG island DNA hypermethylation in 90-95% of prostate cancers. However, prostate cancers expressing GSTP1 have not been well characterized. We used immunohistochemistry against GSTP1 to examine 1673 primary prostatic adenocarcinomas on TMAs with redundant sampling from the index tumor from prostatectomies. GSTP1 protein was positive in at least one TMA core in 7.7% of cases and in all TMA cores in 4.4% of cases. The percentage of adenocarcinomas from Black patients who had any GSTP1 positive TMA cores was 14.9%, which was 2.5 times higher than the percentage from White patients (5.9%; P < 0.001). Further, the percentages of tumors from Black patients who had all TMA spots positive for GSTP1 (9.5%) was 3-fold higher than the percentage from White patients (3.2%; P<0.001). The increased percentage of GSTP1 positive cases in Black men was present only in ERG positive cases. By *in situ* hybridization, GSTP1 mRNA expression was concordant with protein staining, supporting the lack of silencing of at least some *GSTP1* alleles in GSTP1-positive tumor cells. This is the first report revealing that the GSTP1-positive prostate cancer subset is substantially over-represented among prostate cancers from Black compared to White men. This observation should prompt additional studies to determine whether GSTP1 positive cases represent a distinct molecular subtype of prostate cancer and whether GSTP1 expression could provide a biological underpinning for the observed disparate outcomes for Black men.

## Introduction

The pi class glutathione S transferase (encoded by the *GSTP1* gene) is a member of the cytosolic superfamily of glutathione-S-transferases. These phase II detoxification enzymes catalyze the conjugation of glutathione to diverse endogenous and exogenous electrophiles(1). In the prostate, GSTP1 is constitutively expressed at high levels in basal epithelial cells, variably expressed in luminal epithelial cells, and epigenetically silenced in approximately 90 to 95% of adenocarcinomas by somatic hypermethylation of its regulatory CpG island(2–5). Hypermethylation of *GSTP1* appears to occur early in prostatic carcinogenesis since it is already present in approximately 70% of high-grade PIN lesions(6,7), the presumed precursor of most invasive carcinomas, where it is silenced specifically in luminal epithelial cells(7,8). Studies in human prostate cancer cells suggest a role for *GSTP1* as a caretaker gene whose loss increases cell survival in response to protracted oxidative injury(9). In mouse models of carcinogen exposure induced cancers of the lung and skin, Gstp1/2 behaves as a tumor suppressor(10,11). In a mouse model of early prostate cancer development induced by human MYC, Gstp1/2 also functions as a tumor suppressor(12).

GSTP1 acts as a homodimer to catalyze reactions between reactive oxygen species, anti-cancer drugs or carcinogens, and glutathione, sometimes ‘detoxifying’ and at other times ‘toxifying’ the substrates(13). By contrast to silencing *GSTP1*, an induction of expression of GSTP1 protein is used to detect early cancer formation in response to carcinogenic exposure in the rat liver(14). Moreover, expression and overexpression of GSTP1 are common in several human tumor types, where it has been implicated in promoting resistance to a number of chemotherapeutic agents(15). Although GSTP1 expression has been estimated to be maintained in 5-10% of prostate cancer cases, little is known about the molecular and clinical features of these cases and only a few studies have directly asked the question of the frequency of positive GSTP1 Immunohistochemical (IHC) staining in prostate cancer(16,17).

Black men are more likely to develop prostate cancer than White men, and are more likely to have poor outcomes if diagnosed with the disease(18). The reasons for the disparity are multifaceted, including inequities in access to high-quality prostate cancer screening, detection, diagnosis, and treatment. In addition, there appear to be at least some biological differences in disease pathogenesis and malignant progression, potentially based on associations of perceived race with genetic variation, or, as a result of different carcinogenic exposures. For example, when compared to prostate cancers from predominantly White men, prostate cancers from Black men contain fewer *TMPRSS2-ERG* fusions(19–22), fewer *PTEN* deletions(20–23), and more *SPOP* mutations(21,22).

In this study we comprehensively surveyed the frequency of GSTP1 protein expression in human clinical prostate cancers by IHC using tissue microarrays (TMAs). Our goal was to begin to determine whether GSTP1-positive prostate cancer represents a distinct molecular subtype, and, whether the prevalence of GSTP1-positivity differs between Black and White men.

## Methods

### Study Population

The study population consisted of 1673 patients who underwent radical prostatectomy (RRP) at a single center between 1993 and 2019, with an age range from 40 to 75 years old and whose prostatectomy tissue was arrayed across 45 TMA blocks. The cancers encompassed all Gleason grade groups and a spectrum of pathological stages. For IHC staining for GSTP1, we used the following TMA sets. TMA set 1 consists of prostatectomy tissue enriched for different Gleason grade groups and pathological stages from 476 cases operated on between 1997 and 2005 (N= 11 TMA blocks). TMA set 2 consists of prostatectomy tissue from 726 patients operated on between 1993 to 2000 (N = 16 TMA blocks) (24). TMA set 3 consists of prostatectomy tissue from 343 Black and White men, operated on between 1993 and 2019 and matched on Gleason score, pathological stage and date of surgery within 2 years (N=9 TMA blocks) (25). TMA set 4 is a newly designed and consists of prostatectomy tissue from 353 men operated on between 2007 and 2015 from a case-cohort design (N=9 TMA blocks; detailed design to be described in a separate publication). These TMAs were all constructed as described(24,26,27) from the index tumor (highest grade) with a 3-4 fold sampling redundancy. To compare IHC staining and the *in situ* hybridization assay for GSTP1 mRNA, we used a novel TMA (TMA set 5), which consists of prostatectomy tissue from 40 patients operated on less than 1 year before TMA construction in which TMA slides are stored at −20 °C, which improves RNA quality for hybridizations (28).

### IHC Staining

We analytically validated an automated IHC assay against human GSTP1 protein using well-known cell lines and controls and human prostate cancer tissues(8). Cell line FFPE blocks were prepared and utilized as described(29,30). LNCaP cells were negative and DU145 and PC3 prostate cancer cell lines were positive for GSTP1 protein as expected (2). As additional controls for assay specificity, we also observed negative staining using FFPE blocks from *Gstp1/2^-/-^* mouse prostate and liver tissues, and positive staining in the same tissues from mice that were humanized for *GSTP1 (31)*. In a subset of the above TMAs we also performed IHC staining for ERG (rabbit recombinant monoclonal antibody; ABCAM EPR3864, Cambridge, UK) and PTEN (rabbit monoclonal; Cell Signaling Technologies, Danvers, MA) which were performed in an automated assay on the Ventana Roche Discovery using the Discovery HQ+ Amp kit. Genetically validated cell line controls were used for antibody specificity for ERG (VCAP cells are positive and PC3, LnCAP and DU145 are negative) and PTEN (PC3 and LnCAP cells are negative and DU145 cells are positive)(32,33). Immunostaining for p63 was carried out using the mouse monoclonal antibody (clone 4A4, Biocare Medical, Concord,CA, USA) as part of a singleplex stain(34) or as part of a cocktail(35) (PIN4 staining, Biocare Medical) on the Ventana Roche Discovery platform.

### Slide Scanning, Image Management and Scoring

Whole TMA slides were scanned on a Hamamatsu Nanozoomer, and imported into Concentriq (from Proscia). Composite images were exported from Concentriq and imported into and visualized in TMAJ(28). For quality control of IHC staining in the TMA spots, we used the fact that a high fraction of non-neoplastic stromal cells routinely stained positively for GSTP1. We used these stromal cells as internal positive controls such that TMA cores that lacked any staining in tumor cells, and all stromal cells, were regarded as non-evaluable and were not included in the analyses. In another small subset of TMA cores, staining was very weak in intensity in both the tumor cells and stromal cells and these were also considered equivocal in terms of staining quality and were not included. Together, this poor quality staining and uninterpretable staining in weak cases comprised ~ 1 % of tissue cores with carcinoma.

Scanned TMA core images were evaluated for the presence of carcinoma by two pathologists with expertise in prostate cancer (I.V. and A.M.D.). Each TMA core containing carcinoma was evaluated for GSTP1 staining positivity using a two-tiered scoring strategy in which any tumor cell staining above background levels (nuclear, cytoplasmic, or both) were considered positive. All others were considered negative. Cases in which there was positive cancer cell staining for GSTP1 protein were categorized in two ways: any patient who had 1) any TMA spot with some positive tumor cell staining or 2) all TMA spots with some positive tumor cells staining; all other cases were considered GSTP1-negative.

In a subset of cases we also assessed ERG and PTEN to determine co-occurrence of GSTP1 positivity with ERG positivity and PTEN loss. ERG was considered positive for a given patient’s tumor if any cancer cells on any TMA cores with cancer were positive and PTEN was considered lost if any TMA cores contained any tumor cells with complete PTEN absence of staining.

### *In Situ* Hybridization for GSTP1 mRNA

We developed a novel *in situ* hybridization assay for GSTP1 mRNA using ACD RNAscope, which we analytically validated (Probe-Hs-GSTP1 Cat No. 453221, kit version 2.5 manual assay according to manufacturer’s recommendations) using cell lines with known GSTP1 expression status for positive and negative controls as described(8).

### Statistical Analysis

Data were tabulated and statistical tests were performed using Stata 15 for Mac OS. The primary analysis for comparisons between groups used GSTP1 protein positivity status for each patient, either any TMA core positive or all cores positive (Tables 2-8). Tests for differences in proportions for any TMA core or all TMA cores positive for GSTP1 protein between Black and White men, between ERG positive and negative men, and between PTEN absent and PTEN present men were performed with the Pearson Chi2 test. Tests for trends for the proportion of men who were GSTP1 protein positive across Gleason grade groups and pathological stages were performed using the Cuzick’s nonparametric trend test across ordered groups.

## Results

We applied an analytically validated automated IHC assay to evaluate GSTP1 protein expression (8) in a series of TMAs constructed from patients with clinically localized primary prostatic adenocarcinomas. A total of 5,757 TMA cores containing prostatic adenocarcinoma from 1,673 patients were evaluable for GSTP1 protein scoring by IHC. These consisted of 1,197 cores from 336 patients that self-identified as Black and 4,560 TMA cores from 1,337 patients that self-identified as White (**Table 1**).

**Table 1:**
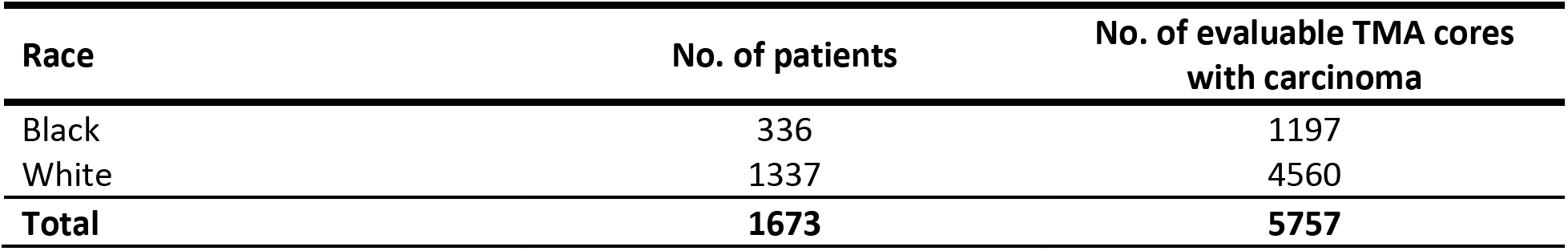
Total number of patients and TMA spots with carcinoma.

| **Race** | **No. of patients** | **No. of evaluable TMA cores**<br>**with carcinoma** |
| --- | --- | --- |
| Black | 336 | 1197 |
| White | 1337 | 4560 |
| **Total** | **1673** | **5757** |

As expected, carcinoma cells in most prostatic adenocarcinomas were completely negative for GSTP1 protein (92.3%; **Table 2**). In a subset of cases, however, some or all cancer cells stained positively for GSTP1. While some cases were uniformly positive for GSTP1, in other cases containing GSTP1 positive tumor cells, we often observed heterogeneity in staining in terms of both cell number and staining intensity. This variability occurred among cancer cells at times within a given TMA core, between a man’s TMA cores and between men. **Figure 1** demonstrates representative images of these patterns of staining. Overall, 7.7% of cases contained any TMA cores, and 4.5% had all TMA cores that contained GSTP1 positive cancer cells.

**Table 2:**
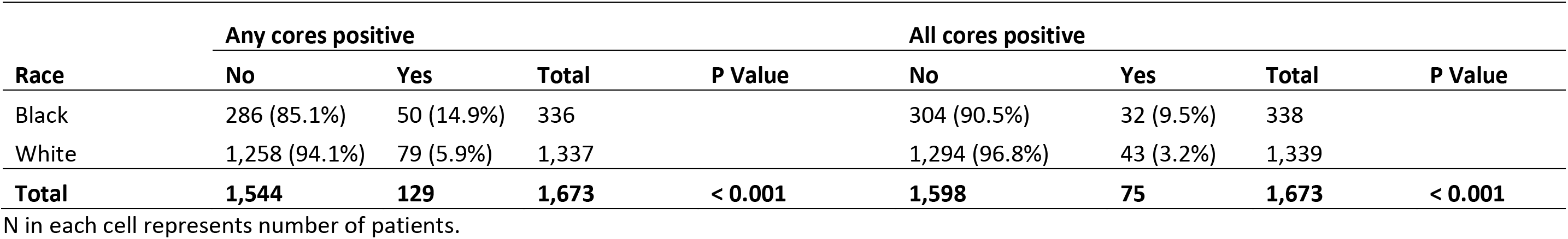
GSTP1 Protein Staining in Cancer Cells in Black and White Patients.

**Figure 1.**
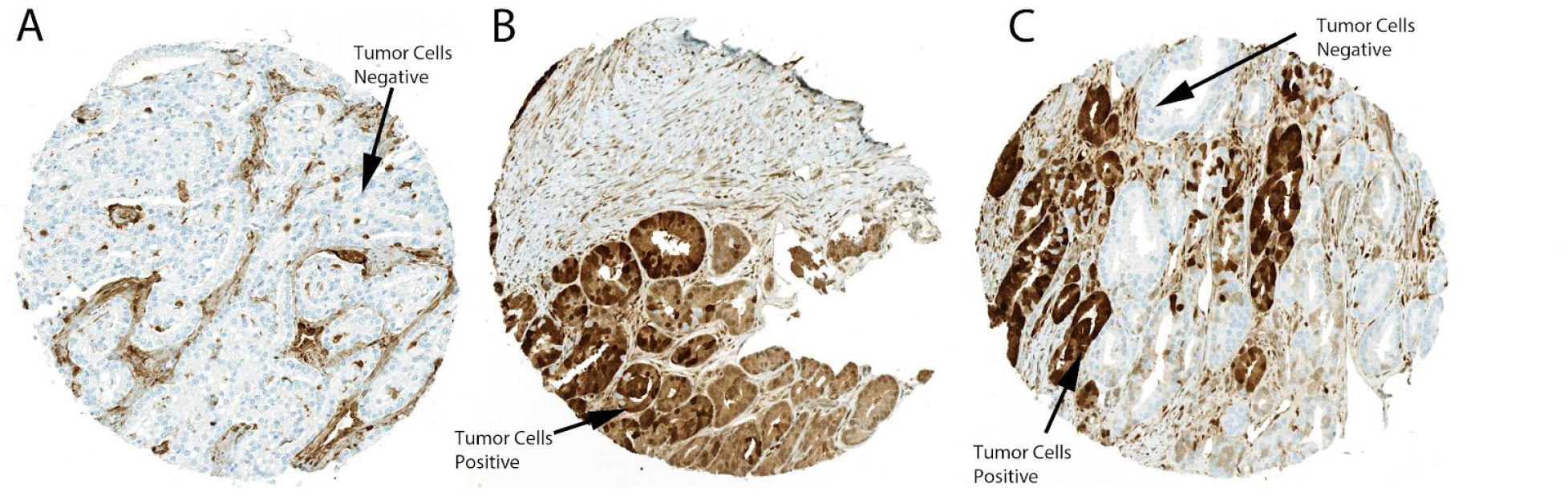
Patterns of GSTP1 protein staining. Medium power views of TMA spots containing adenocarcinoma with examples of all tumor cells being GSTPI-negative (A), all tumor cells being GSTPI-positive (B), and a spot with heterogeneous mosaic staining with some cells staining positively and some negatively (C). In all examples, there is abundant stromal cell staining that is most apparent in (A) in between tumor nests. Original magnification x 100.

The percentage of adenocarcinomas from Black patients who had any GSTP1 positive TMA cores was 14.9%, which was 2.5 times higher than the percentage of cases with any GSTP1 positive cores from White patients, which was statistically significant (P < 0.001) (**Table 2**). Further, the percentages of tumors from Black patients who had all TMA spots with cancer cells staining positive for GSTP1 (9.5%) was 3-fold higher than the percentage of tumors from White patients (3.2%), which was also statistically significant (P<0.001) (**Table 2**). We confirmed that the number of evaluable TMA cores with carcinoma was similar among tumors from Black and White men, and also ran sensitivity analysis restricted to men with the same number of evaluable cores and similar results were obtained (not shown).

Cases with any or all TMA cores with cancer cells staining positive for GSTP1 occurred across all Gleason grade groups and stages, and the higher percentage of cases from Black patients positive for GSTP1 was apparent among all grade groups except in grade group 5 (**Table 3**) and except in stage group 3 (**Table 4**). In Black but not White patients, the percentage of cases with any core positive decreased with increasing Gleason grade group (**Table 3**) and with increasing pathological stage (**Table 4**), but these trends were not statistically significant.

**Table 3.** GSTP1 Protein Staining by Gleason Grade Groups in Black and White Patients.

| Grade group | Black |  |  |  | White |  |  |  |
| --- | --- | --- | --- | --- | --- | --- | --- | --- |
|  | <u>Any cores positive</u> |  |  | P Value | <u>Any cores positive</u> |  |  | P Value |
|  | No | Yes | Total |  | No | Yes | Total |  |
| 1 (GS 6) | 68 (77.3%) | 20 (22.7%) | 88 (100%) |  | 230 (93.1%) | 17 (6.9%) | 247 (100%) |  |
| 2 (GS 3+4=7) | 97 (86.6%) | 15 (13.4%) | 112 (100%) |  | 448 (94.7) | 25 (5.3%) | 473 (100%) |  |
| 3 (GS 4+3=7) | 43 (89.6%) | 5 (10.4%) | 48 (100%) |  | 287 (95.7%) | 13 (4.3%) | 300 (100%) |  |
| 4 (GS 8) | 24 (82.8%) | 5 (17.2%) | 29 (100%) |  | 150 (93.2%) | 11 (6.8%) | 161 (100%) |  |
| 5 (GS 9-10) | 54 (91.5%) | 5 (8.5%) | 59 (100%) |  | 143 (91.7%) | 13 (8.3%) | 156 (100%) |  |
| **Total** | **286 (84.9%)** | **50 (14.8%)** | **336 (100%)** | **0.039** | **1,258 (94.0%)** | **79 (5.9%)** | **1,337 (100%)** | **0.50** |

| Grade group | <u>All cores positive</u> |  |  |  | <u>All cores positive</u> |  |  |  |
| --- | --- | --- | --- | --- | --- | --- | --- | --- |
|  | No | Yes | Total | P Value | No | Yes | Total | P Value |
| 1 (GS 6) | 74 (84%) | 14 (15.9%) | 88 (100%) |  | 238 (96.3%) | 9 (3.6%) | 247 (100%) |  |
| 2 (GS 3+4=7) | 106 (94.6%) | 6 (5.4%) | 112 (100%) |  | 460 (97.3%) | 13 (2.8%) | 473 (100%) |  |
| 3 (GS 4+3=7) | 44 (91.7%) | 4 (8.3%) | 48 (100%) |  | 294 (98.0%) | 6 (2%) | 300 (100%) |  |
| 4 (GS 8) | 24 (82.8%) | 5 (17.2%) | 29 (100%) |  | 152 (94.4%) | 9 (5.6%) | 161 (100%) |  |
| 5 (GS 9-10) | 56 (94.9%) | 3 (5.1%) | 61 (100%) |  | 150 (96.2%) | 6 (3.9%) | 156 (100%) |  |
| **Total** | **304 (90.5%)** | **32 (9.5%)** | **338 (100%)** | **0.21** | **1294 (96.8%)** | **43 (3.2%)** | **1,337 (100%)** | **0.46** |

N in each cell represents number of patients.

**Table 4.** GSTP1 Protein Staining by Pathological Stage in Black and White Patients.

| Stage | Black |  |  |  | White |  |  |  |
| --- | --- | --- | --- | --- | --- | --- | --- | --- |
|  | <u>Any cores positive</u> |  | Total | P Value | <u>Any cores positive</u> |  | Total | P Value |
|  | No | Yes |  |  | No | Yes |  |  |
| 1 | 129 (78.7%) | 35 (21.3%) | 164 |  | 459 (93.1%) | 34 (6.9%) | 493 |  |
| 2 | 87 (89.7%) | 10 (10.3%) | 97 |  | 507 (96.0%) | 21 (4.0%) | 528 |  |
| 3 | 69 (93.2%) | 5 (6.8%) | 74 |  | 287 (92.3%) | 24 (7.7%) | 311 |  |
| **Total** | **285 (85.1%)** | **50 (14.9%)** | **335** | **<0.004** | **1253 (94%)** | **79 (5.9%)** | **1332** | **0.045** |

| Stage | <u>All cores positive</u> |  |  |  | <u>All cores positive</u> |  |  |  |
| --- | --- | --- | --- | --- | --- | --- | --- | --- |
|  | No | Yes | Total | P Value | No | Yes | Total | P Value |
| 1 | 143 (87.2%) | 21 (12.8%) | 164 |  | 476 (96.6%) | 17 (3.5%) | 493 |  |
| 2 | 91 (93.8%) | 6 (6.2%) | 97 |  | 515 (97.5%) | 13 (2.5%) | 528 |  |
| 3 | 69 (93.2%) | 5 (6.8%) | 74 |  | 298 (95.8%) | 13 (4.2%) | 311 |  |
| **Total** | **303 (90.5%)** | **32 (9.5%)** | **335** | **0.139** | **1289 (96.8%)** | **43 (3.2%)** | **1332** | **0.373** |

Stage groups: 1= "Organ Confined" - T2N0; 2 – "Extraprostatic Extension" - T3aN0; 3 = Seminal Vesicle Invasion or Pelvic Lymph Node Metastasis - T3bN0 or any N1 (AJCC 2007 staging criteria). N in each cell represents number of patients.

A subset of TMAs (N= 16 TMA blocks) were stained for ERG protein by IHC, which tightly correlates with *TMPRSS2-ERG* gene fusions(36,37). *TMPRSS2-ERG* gene fusions are the most common form of *ETS* gene family member fusions in prostate cancer and are mutually exclusive with other *ETS* family member fusions and a number of other somatic molecular alterations in prostate cancer, such as *SPOP* mutations or SPINK overexpression(38). Therefore, prostate cancers harboring *ERG* gene fusions are considered a distinct molecular subtype of prostate cancer. In this TMA subset, the percentage of tumors from Black patients with any TMA core positive for ERG was significantly lower than for tumors from Whilte patients, as seen by many groups previously(19) (**Table 6**).

When not considering race, the percentage of cases with any TMA spot or all TMA spots with cancer cells staining positive for GSTP1 was lower in those who were ERG+ than ERG- (**Table 5**). This same pattern was observed when examining White patients only (**Table 6**, any TMA spot positive). In Black patients, the opposite pattern was observed; the percentage of any TMA spot positive for GSTP1 was higher in men who were ERG positive than ERG negative. The percentage of patients who were both ERG positive and GSTP1 positive was 23.1% in Black men, but only 5.1% in White men, a difference that was statistically significant (**Table 6**, P<0.001). In contrast, the percentage of patients who were ERG negative and GSTP1 positive was more similar in Black (12.9%) and White (9.4%) patients (**Table 6**, P=0.38).

**Table 5.**
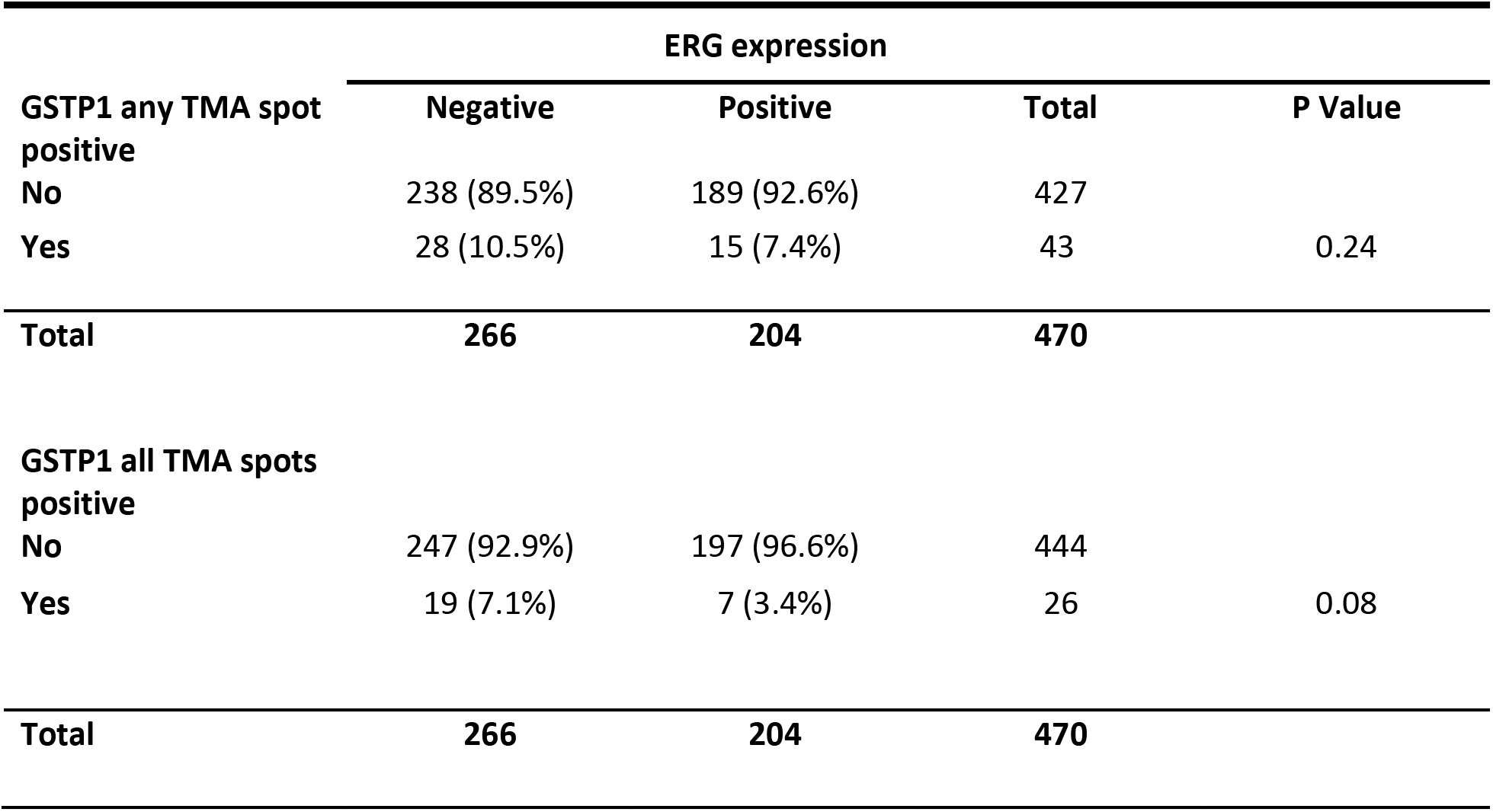
GSTP1 Protein Staining by ERG Status Overall.

**Table 6.**
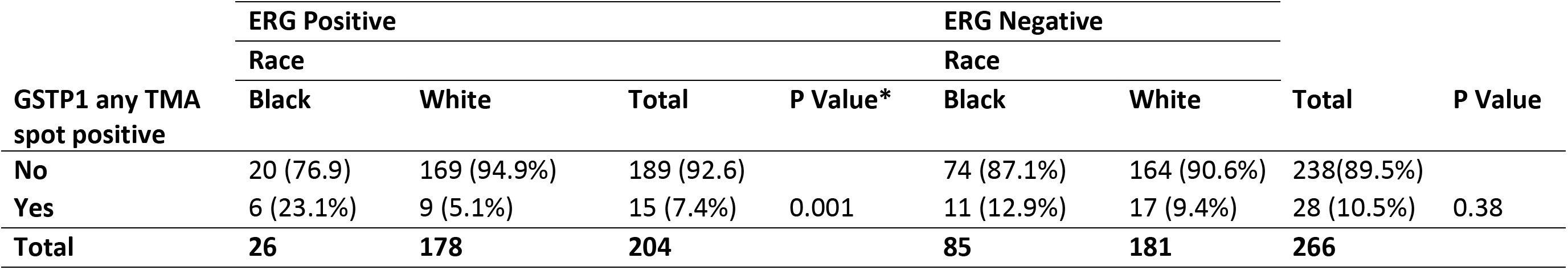
GSTP1 Protein Staining by ERG Status in Black and White patients.

A subset of TMAs (N=16) were also stained by IHC for PTEN, which is a known tumor suppressor in prostate cancer, whose loss is associated with disease progression(23). Prior work has also found a lower fraction of prostatic carcinomas from Black patients with PTEN loss than carcinomas from White patients (20,21), which we observed here (**Table 8**; 22.5% PTEN loss for Black patients and 34.9% PTEN loss for White patients). When not considering race, the percentage of patients with any GSTP1-positive TMA spot appeared to be lower in men with any PTEN loss than with PTEN intact (6.6% compared with 10.3%), but this difference was not statistically significant (**Table 7**). However, stratifying by race, the percentage of any TMA spot with cells staining positive for GSTP1 was comparable in those with PTEN loss and PTEN intact in Black men and in White men (**Table 8**). The percentage of patients who had both PTEN loss and were GSTP1 positive was 12.0% in Black men, but only 5.6% in White men; this difference was not statistically significant (**Table 8**, P=0.24). These percentages were similar to those in those who had both PTEN intact and were GSTP1 positive, with 16.5% in Black men and 8.1% in White men; this difference was statistically significant (**Table 8**, P=0.029).

**Table 7.** GSTP1 Protein Staining by PTEN Loss Overall.

| **GSTP1 any TMA spot**<br>**positive** | **PTEN status** |  | **Total** |
| --- | --- | --- | --- |
|  | **Loss (any)** | **Intact** |  |
| **No** | 141 (93.4%) | 287 (89.7%) | 428 |
| **Yes** | 10 (6.6%) | 33 (10.3%) | 43 |
| **Total** | **151** | **320** | **471** |

P = 0.19

**Table 8.**
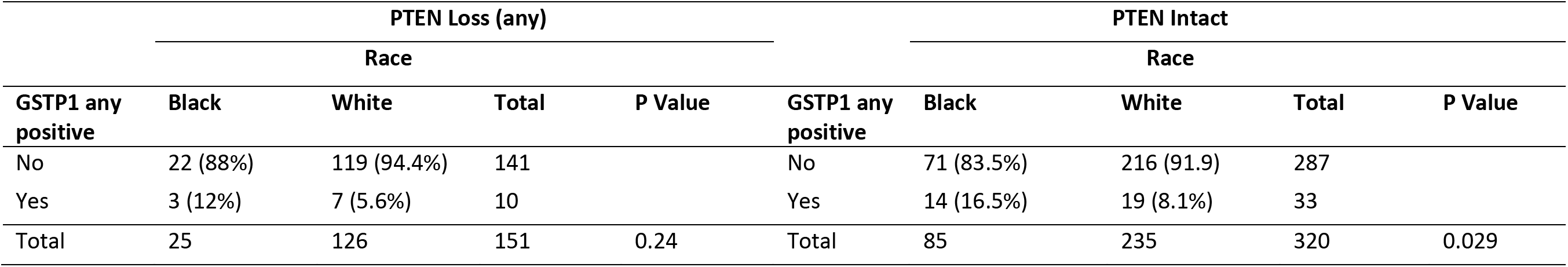
GSTP1 Protein Staining by PTEN Loss in Black and White Patients.

Rare cases of prostate cancer have been reported that are strongly positive for nuclear p63 staining (34,39–41), and these cases tend to express GSTP1, and at times lack GSTP1 GpG island methylation(34). We asked whether GSTP1 positive cases in the current study were ever positive for p63 nuclear staining. P63 IHC staining was available for 22 of the cases across 6 of the TMA blocks that stained positively for GSTP1 that have also been stained in our laboratory for p63, and none of these cases (0/22) were positive for nuclear p63 staining. We conclude that, while many p63-positive cancers are also positive for GSTP1(34), most GSTP1-positive cancers are not p63-positive.

To test whether samples with GSTP1 positive protein staining also contained GSTP1 mRNA expression, we developed an ACD RNAscope *in situ* hybridization assay. We tested the hybridization reaction on cell lines and found that, as expected, PC3 cells and DU145 cells were positive and LNCaP cells were negative(2) (**Figure 2**). In normal-appearing human prostate tissue, GSTP1 protein is consistently highly expressed in basal cells, with much lower and variable expression in normal appearing luminal cells. Furthermore, the expression is highly elevated in many of the intermediate luminal cells present in the atrophic epithelium of proliferative inflammatory atrophy(7,42). Using standard slides from RRP specimens we found a similar pattern of hybridization signals for the GSTP1 ACD RNAscope probe set. There were also strong signals found in regions of the urethra as well as ejaculatory ducts, which is consistent with prior studies on GSTP1 protein(43) (**Figure 2**). Since RNAscope assays perform less robustly on older prostate cancer specimens(28), we performed *in situ* hybridization for GSTP1 mRNA and IHC using a “recent case” TMA consisting of an array from 40 RRP cases (TMA set 5). We found 4 cases positive for GSTP1 protein by IHC and the same cases were also positive for GSTP1 mRNA; all cases staining negatively for GSTP1 protein were also negative for GSTP1 mRNA, giving a 100% concordance (data not shown). We also performed IHC and *in situ* hybridization for GSTP1 mRNA on a number of standard slides from prostatectomy specimens and found an example that was heterogeneous for GSTP1 protein in the tumor. **Figure 3** shows a region from this case that shows tight concordance between GSTP1 protein and mRNA expression.

**Figure 2.**
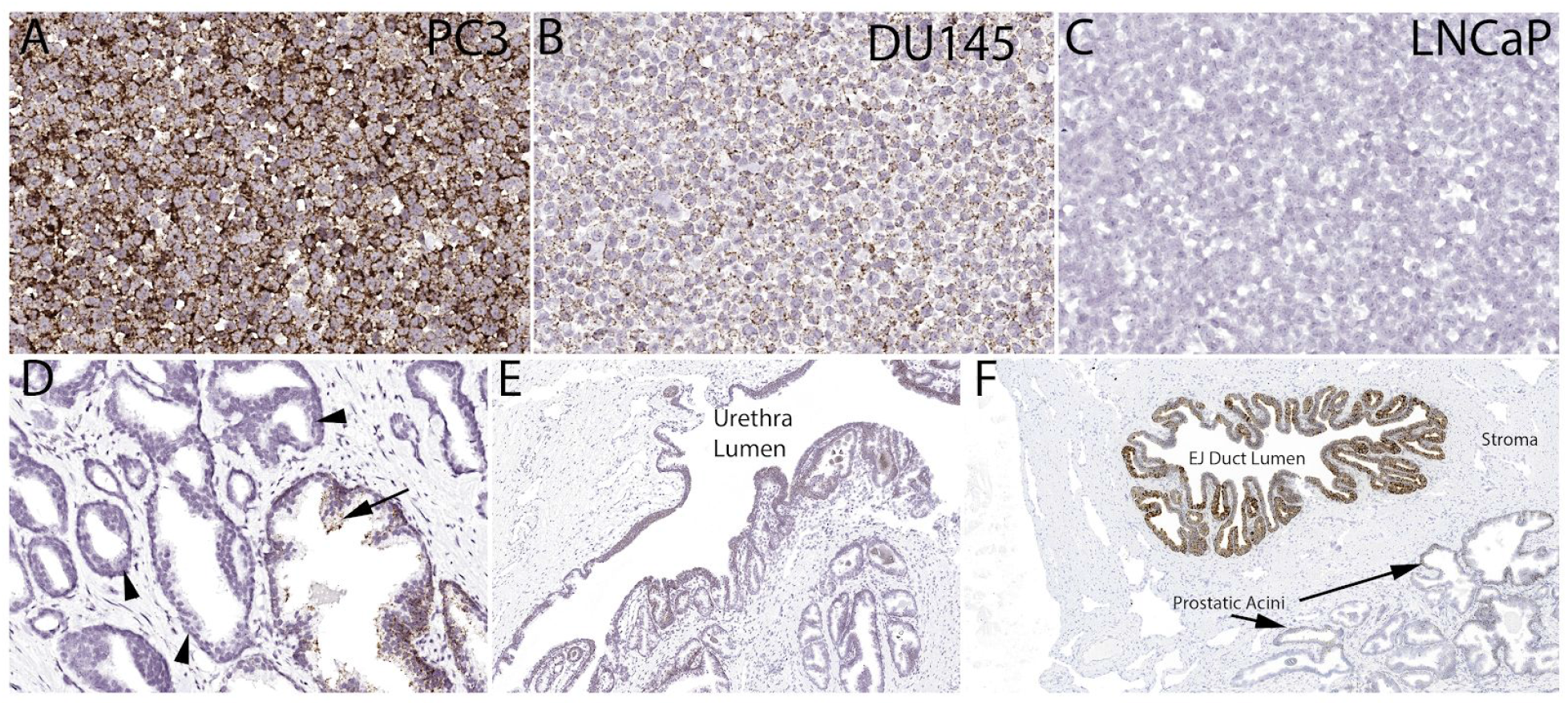
*In situ* hybridization for GSTP1 mRNA in cell lines (A-C) and benign prostate (D-F). EJ duct lumen indicates ejaculatory duct lumen.

**Figure 3.**
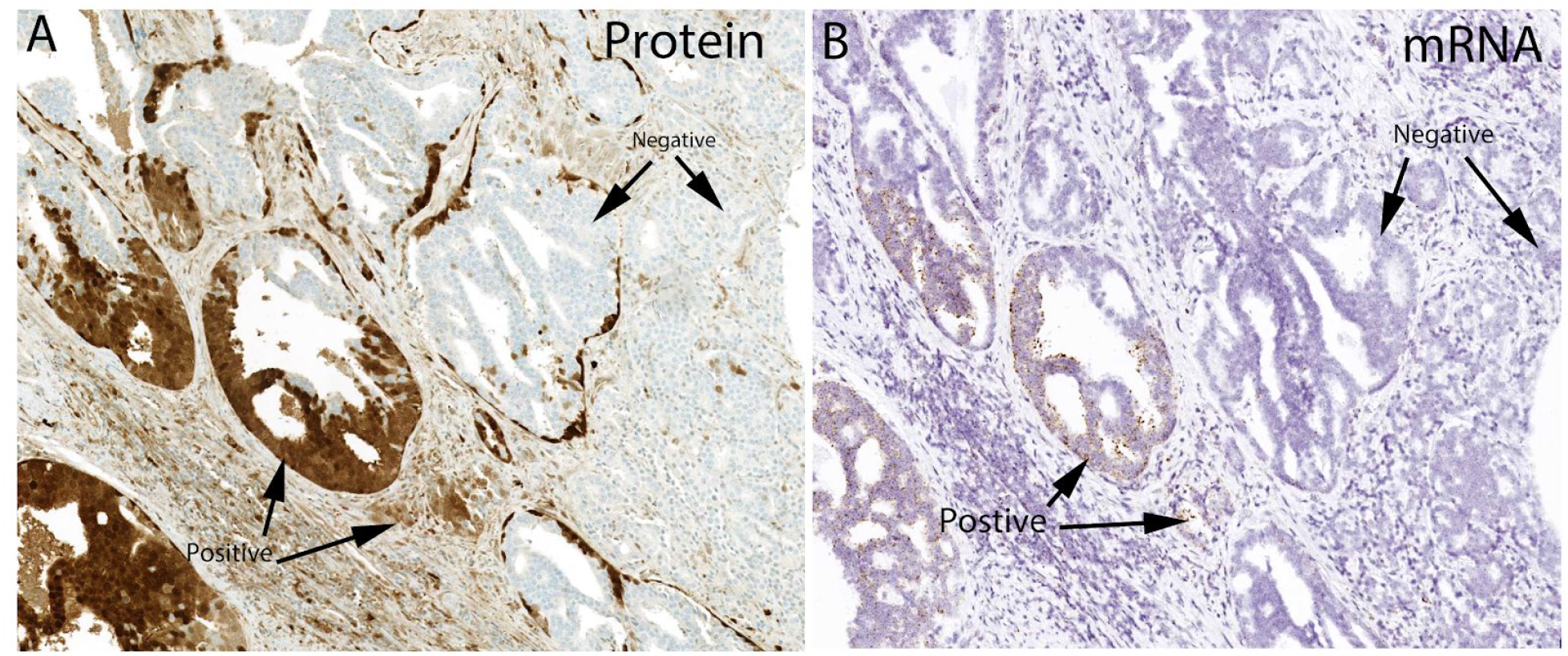
Concordance between *in situ* hybridization and IHC for GSTP1 in a heterogeneous prostate cancer case.

## Discussion

In this study, we used a large number of primary untreated prostate cancer cases from prostatectomies (N=1673) to estimate the percentage of prostatic adenocarcinomas that are positive for GSTP1 protein by IHC. We found an overall rate of any positive staining in cancer foci of 7.7%. Interestingly, we also found that GSTP1-positive carcinomas were often heterogeneous for GSTP1 protein (41%). *In situ* hybridization for GSTP1 mRNA showed that the increased protein in the GSTP1-positive cases likely occurred through transcriptional upregulation, suggested by the tight correlation between the protein and mRNA levels. For the first time, we report that GSTP1-positive cases are significantly more common in prostatic adenocarcinomas from Black patients compared with White patients (2.5-3 fold).

Given the difference in the percentage of GSTP1 positive cases by race, the current findings raise the question of whether GSTP1 positive cases represent a distinct molecular subtype of this disease. Interestingly, we found that the increase in GSTP1-positive cancers in Black men was greater in ERG-positive cases. This is of interest because prostate adenocarcinomas from Black men harbor ERG gene rearrangements less frequently than those from White men. In terms of PTEN, there was a similar increase in the percentage of GSTP1-positive cases by PTEN loss and PTEN intact in Black patients.

Genomics studies of prostate cancers over the last several years have revealed the long tail of molecular alterations that can lead to cancer and influence disease progression and drug resistance (44–49). These long tail events include low frequency germline and/or somatic molecular alterations involved in DNA repair (e.g. MMR defects caused by 1 of 4 different genes, BRCA2, ATM and CDK12). Other low frequency events considered important include FOXA1 mutations, IDH1/2 mutations, and mutations in a number of genes that affect epigenetic chromatin modifications. Given the large breadth of molecular alterations in prostate cancer, additional studies with larger numbers of patients are required to determine whether GSTP1 positive cases represent a distinct molecular subtype of prostate cancer.

In terms of disease aggressiveness, the percentage of men whose tumors were GSTP1-positive tended to decrease with increasing grade and stage in Black men, but not in White men (**Tables 3-4**). Thus, the increased frequency of GSTP1-positive cancers in Black versus Whilte men occurred in low and intermediate grade and stage cancers only (**Tables 3-4**).

This study was focused on whether GSTP1-positive cases are a distinct molecular subtype and on Black/White differences in GSTP1 positivity. In terms of patient outcomes, in a series of 640 cases stained by IHC, there was no prognostic significance in univariate analysis for GSTP1 status(16). Additional studies are underway to determine whether GSTP1 positive cases differ in terms of rates of biochemical recurrence, metastatic disease and deaths due to prostate cancer. Further, it will be of interest to determine whether GSTP1 status is related to response to therapies such as androgen deprivation, radiation, or chemotherapy and whether there are differences in the percentage of cases that are GSTP1 positive in metastatic castration sensitive and castration resistant prostate cancers.

In terms of prostate cancer etiology, major risk factors are ancestry, family history and age. In terms of environmental exposures at present, the fraction of prostate cancers attributable to known exposures remains quite low(50). At present, we have not determined whether men with prostate cancer that are GSTP1 positive, whether they be from Black or White men, have had different exposures than men with prostate cancers that are GSTP1 negative. Additional studies from population-based cohort studies with extensive pre-diagnosis exposure information are required to address this important issue and the potential racial disparity in prostate cancer burden.

To determine whether there is a different etiology for prostate cancers developing in Black versus Whte patients, it may be instructive to look at gene expression studies. Interestingly, when probing for differences in gene expression in prostate cancers from Black men and White men, a number of studies(21,51–53), albeit not all(54), have reported increases in inflammatory cytokine signaling in tumors from men of African descent. While inflammatory signaling and altered inflammatory cell infiltrates in the tumor microenvironment may occur during any and all temporal phases of cancer development and progression, we have previously postulated that chronic long standing inflammation may lead to prostate cancer most likely via lesions we termed proliferative inflammatory atrophy(42,55,56). Interestingly, in regions of PIA, there appears to be an induction of GSTP1 in luminal cells, which we refer to as intermediate luminal cells (42,56,57); with progression to PIN or adenocarcinoma accompanied by CpG island hypermethylation and loss of expression (7). In terms of GSTP1 positive prostate cancer, it is possible that some cases arise after induction of GSTP1 in luminal cells and without silencing the gene. Martignano et al. found a strong correlation with *GSTP1* CpG island hypermethylation and GSTP1 protein status by IHC (17). Additional studies are underway in our laboratory to determine the CpG methylation status of *GSTP1* alleles in cases with positive staining.

Finally, the finding that GSTP1-positive prostate cancer subset is substantially over-represented among prostate cancers from Black compared White men could be of importance from the standpoint of treatment of lethal prostate cancer, providing a potential biological underpinning, at least in part, for the observed disparate outcomes for Black men. In breast cancer, the presence or absence of GSTP1 expression predicts response to cytotoxic chemotherapy, particularly to the taxanes docetaxel and paclitaxel(58–61). Forced expression of the enzyme in GSTP1-negative breast cancer cells confers docetaxel resistance(58). In addition, an MCF-7 clone selected for taxane-resistance exhibited activation of GSTP1 expression(62). Thus, the over-representation of GSTP1-positive prostate cancer in Black men may render them less responsive to taxane chemotherapy, which, besides androgen deprivation therapy, is the preferred therapy for advanced, lethal metastatic prostate cancer(5).

In summary, we systematically examined GSTP1 protein expression in a large number of primary prostatic adenocarcinomas and found an overall rate of positivity of 7.7%, and a higher percentage of cases staining positive in Black men compared with cases from White men (2.5-3 fold). A significant difference in the percentages of GSP1 positive cases in Black men was present only in ERG-positive cases, as compared with ERG-negative caes. These findings should stimulate additional studies regarding whether GSTP1 positive cases are a unique molecular and clinico-pathological subtype of human prostate cancer, with potential implications for disease etiology, racial disparities and therapeutic resistance mechanisms.

## Data Availability

not applicable

## Acknowledgements

Funding: Supported by NIH/NCI SPORE in Prostate Cancer: P50CA58236, and the NIH/NCI U01 CA196390 for the Molecular and Cellular Characterization of Screen Detected Lesions (MCL), the U.S. Department of Defense Prostate Cancer Research Program (PCRP): W81XWH-18-2-0015. The Johns Hopkins Sidney Kimmel Comprehensive Cancer Center Oncology Tissue Services Laboratory supported by NIH/NCI grant P30 CA006973.

## References

1. Allocati N, Masulli M, Di Ilio C, Federici L. Glutathione transferases: substrates, inihibitors and pro-drugs in cancer and neurodegenerative diseases. Oncogenesis [Internet]. 2018;7:8. Available from: http://dx.doi.org/10.1038/s41389-017-0025-3

2. Lee WH, Morton RA, Epstein JI, Brooks JD, Campbell PA, Bova GS, et al. Cytidine methylation of regulatory sequences near the pi-class glutathione S-transferase gene accompanies human prostatic carcinogenesis. Proc Natl Acad Sci U S A [Internet]. 1994;91:11733–7. Available from: https://www.ncbi.nlm.nih.gov/pubmed/7972132

3. Lin X, Tascilar M, Lee WH, Vles WJ, Lee BH, Veeraswamy R, et al. GSTP1 CpG island hypermethylation is responsible for the absence of GSTP1 expression in human prostate cancer cells. Am J Pathol [Internet]. 2001;159:1815–26. Available from: http://dx.doi.org/10.1016/S0002-9440(10)63028-3

4. Yegnasubramanian S, De Marzo AM, Nelson WG. Prostate Cancer Epigenetics: From Basic Mechanisms to Clinical Implications. Cold Spring Harb Perspect Med [Internet]. 2018; Available from: http://dx.doi.org/10.1101/cshperspect.a030445

5. Nelson WG, Antonarakis ES, Balentine CH, De Marzo AM, Deweese TL. Prostate Cancer. Abeloff’s Clinical Oncology, Sixth Edition [Internet]. Elsevier, Inc; 2020 [cited 2020 Mar 31]. page 1401–32. Available from: https://www-clinicalkey-com.proxy1.library.jhu.edu/#!/content/book/3-s2.0-B9780323476744000815

6. Brooks JD, Weinstein M, Lin X, Sun Y, Pin SS, Bova GS, et al. CG island methylation changes near the GSTP1 gene in prostatic intraepithelial neoplasia. Cancer Epidemiol Biomarkers Prev [Internet]. 1998;7:531–6. Available from: https://www.ncbi.nlm.nih.gov/pubmed/9641498

7. Nakayama M, Bennett CJ, Hicks JL, Epstein JI, Platz EA, Nelson WG, et al. Hypermethylation of the human glutathione S-transferase-pi gene (GSTP1) CpG island is present in a subset of proliferative inflammatory atrophy lesions but not in normal or hyperplastic epithelium of the prostate: a detailed study using laser-capture microdissection. Am J Pathol [Internet]. 2003;163:923–33. Available from: https://www.ncbi.nlm.nih.gov/pubmed/12937133

8. Trabzonlu L, Kulac I, Zheng Q, Hicks JL, Haffner MC, Nelson WG, et al. Molecular Pathology of High-Grade Prostatic Intraepithelial Neoplasia: Challenges and Opportunities. Cold Spring Harb Perspect Med [Internet]. 2018; Available from: http://dx.doi.org/10.1101/cshperspect.a030403

9. Mian OY, Khattab MH, Hedayati M, Coulter J, Abubaker-Sharif B, Schwaninger JM, et al. GSTP1 Loss results in accumulation of oxidative DNA base damage and promotes prostate cancer cell survival following exposure to protracted oxidative stress. Prostate [Internet]. 2016;76:199–206. Available from: http://dx.doi.org/10.1002/pros.23111

10. Henderson CJ, Smith AG, Ure J, Brown K, Bacon EJ, Wolf CR. Increased skin tumorigenesis in mice lacking pi class glutathione S-transferases. Proc Natl Acad Sci U S A [Internet]. 1998;95:5275–80. Available from: https://www.ncbi.nlm.nih.gov/pubmed/9560266

11. Ritchie KJ, Henderson CJ, Wang XJ, Vassieva O, Carrie D, Farmer PB, et al. Glutathione transferase pi plays a critical role in the development of lung carcinogenesis following exposure to tobacco-related carcinogens and urethane. Cancer Res [Internet]. 2007;67:9248–57. Available from: http://dx.doi.org/10.1158/0008-5472.CAN-07-1764

12. Iwata T, Schultz D, Vaughn M, Yegnasubramanian S, Nelson WG, De Marzo AM. Glutathione S-transferase Pi (gstp1) Deficiency Accelerates Prostate Carcinogenesis In The Lo-myc Mouse. J Urol. Elsevier; 2009;181:183–4.

13. Townsend DM, Tew KD. The role of glutathione-S-transferase in anti-cancer drug resistance. Oncogene [Internet]. 2003;22:7369–75. Available from: http://dx.doi.org/10.1038/sj.onc.1206940

14. Satoh K, Kitahara A, Soma Y, Inaba Y, Hatayama I, Sato K. Purification, induction, and distribution of placental glutathione transferase: a new marker enzyme for preneoplastic cells in the rat chemical hepatocarcinogenesis. Proc Natl Acad Sci U S A [Internet]. 1985;82:3964–8. Available from: http://dx.doi.org/10.1073/pnas.82.12.3964

15. Hayes JD, Pulford DJ. The glutathione S-transferase supergene family: regulation of GST and the contribution of the isoenzymes to cancer chemoprotection and drug resistance. Crit Rev Biochem Mol Biol [Internet]. 1995;30:445–600. Available from: http://dx.doi.org/10.3109/10409239509083491

16. Sailer V, Eberhard HLK, Stephan C, Wernert N, Perner S, Jung K, et al. Glutathione S-transferase-pi protein expression in prostate cancer--not always a useful diagnostic tool. Histopathology [Internet]. 2015;67:577–9. Available from: http://dx.doi.org/10.1111/his.12671

17. Martignano F, Gurioli G, Salvi S, Calistri D, Costantini M, Gunelli R, et al. GSTP1 Methylation and Protein Expression in Prostate Cancer: Diagnostic Implications. Dis Markers [Internet]. 2016;2016:4358292. Available from: http://dx.doi.org/10.1155/2016/4358292

18. Smith ZL, Eggener SE, Murphy AB. African-American Prostate Cancer Disparities. Curr Urol Rep [Internet]. 2017;18:81. Available from: http://dx.doi.org/10.1007/s11934-017-0724-5

19. Rosen P, Pfister D, Young D, Petrovics G, Chen Y, Cullen J, et al. Differences in frequency of ERG oncoprotein expression between index tumors of Caucasian and African American patients with prostate cancer. Urology [Internet]. 2012;80:749–53. Available from: http://dx.doi.org/10.1016/j.urology.2012.07.001

20. Tosoian JJ, Almutairi F, Morais CL, Glavaris S, Hicks J, Sundi D, et al. Prevalence and Prognostic Significance of PTEN Loss in African-American and European-American Men Undergoing Radical Prostatectomy. Eur Urol [Internet]. 2017;71:697–700. Available from: http://dx.doi.Org/10.1016/j.eururo.2016.07.026

21. Yuan J, Kensler KH, Hu Z, Zhang Y, Zhang T, Jiang J, et al. Integrative comparison of the genomic and transcriptomic landscape between prostate cancer patients of predominantly African or European genetic ancestry. PLoS Genet [Internet]. 2020;16:e1008641. Available from: http://dx.doi.org/10.1371/journal.pgen.1008641

22. Koga Y, Song H, Chalmers ZR, Newberg J, Kim E, Carrot-Zhang J, et al. Genomic Profiling of Prostate Cancers from Men with African and European Ancestry. Clin Cancer Res [Internet]. 2020; Available from: http://dx.doi.org/10.1158/1078-0432.CCR-19-4112

23. Jamaspishvili T, Berman DM, Ross AE, Scher HI, De Marzo AM, Squire JA, et al. Clinical implications of PTEN loss in prostate cancer. Nat Rev Urol [Internet]. 2018; Available from: http://dx.doi.org/10.1038/nrurol.2018.9

24. Chaux A, Peskoe SB, Gonzalez-Roibon N, Schultz L, Albadine R, Hicks J, et al. Loss of PTEN expression is associated with increased risk of recurrence after prostatectomy for clinically localized prostate cancer. Mod Pathol [Internet]. 2012;25:1543–9. Available from: http://dx.doi.org/10.1038/modpathol.2012.104

25. Hempel Sullivan H, Heaphy CM, Kulac I, Cuka N, Lu J, Barber JR, et al. High Extratumoral Mast Cell Counts Are Associated with a Higher Risk of Adverse Prostate Cancer Outcomes. Cancer Epidemiol Biomarkers Prev [Internet]. 2020; Available from: http://dx.doi.org/10.1158/1055-9965.EPI-19-0962

26. Faith DA, Isaacs WB, Morgan JD, Fedor HL, Hicks JL, Mangold LA, et al. Trefoil factor 3 overexpression in prostatic carcinoma: prognostic importance using tissue microarrays. Prostate [Internet]. Wiley Online Library; 2004;61:215–27. Available from: http://dx.doi.org/10.1002/pros.20095

27. Gurel B, Iwata T, Koh CM, Jenkins RB, Lan F, Van Dang C, et al. Nuclear MYC protein overexpression is an early alteration in human prostate carcinogenesis. Mod Pathol [Internet]. United States and Canadian Academy of Pathology, Inc.; 2008;21:1156–67. Available from: http://dx.doi.org/10.1038/modpathol.2008.111

28. Baena-Del Valle JA, Zheng Q, Hicks JL, Fedor H, Trock BJ, Morrissey C, et al. Rapid Loss of RNA Detection by In Situ Hybridization in Stored Tissue Blocks and Preservation by Cold Storage of Unstained Slides. Am J Clin Pathol [Internet]. 2017 [cited 2017 Oct 18];148:398–415. Available from: http://dx.doi.org/10.1093/ajcp/aqx094

29. Holdhoff M, Guner G, Rodriguez FJ, Hicks JL, Zheng Q, Forman MS, et al. Absence of Cytomegalovirus in Glioblastoma and Other High-grade Gliomas by Real-time PCR, Immunohistochemistry, and In Situ Hybridization. Clin Cancer Res [Internet]. 2017;23:3150–7. Available from: http://dx.doi.org/10.1158/1078-0432.CCR-16-1490

30. Chen J, Zheng Q, Peiffer LB, Hicks JL, Haffner MC, Rosenberg AZ, et al. An in situ atlas of mitochondrial DNA in mammalian tissues reveals high content in stem/progenitor cells. Am J Pathol [Internet]. 2020; Available from: http://dx.doi.org/10.1016/j.ajpath.2020.03.018

31. Vaughn MP, Biswal Shinohara D, Castagna N, Hicks JL, Netto G, De Marzo AM, et al. Humanizing n-class glutathione S-transferase regulation in a mouse model alters liver toxicity in response to acetaminophen overdose. PLoS One [Internet]. 2011;6:e25707. Available from: http://dx.doi.org/10.1371/journal.pone.0025707

32. Lotan TL, Gurel B, Sutcliffe S, Esopi D, Liu W, Xu J, et al. PTEN protein loss by immunostaining: analytic validation and prognostic indicator for a high risk surgical cohort of prostate cancer patients. Clin Cancer Res [Internet]. 2011;17:6563–73. Available from: http://dx.doi.org/10.1158/1078-0432.CCR-11-1244

33. Toubaji A, Albadine R, Meeker AK, Isaacs WB, Lotan T, Haffner MC, et al. Increased gene copy number of ERG on chromosome 21 but not TMPRSS2-ERG fusion predicts outcome in prostatic adenocarcinomas. Mod Pathol [Internet]. United States and Canadian Academy of Pathology, Inc.; 2011;24:1511–20. Available from: http://dx.doi.org/10.1038/modpathol.2011.111

34. Tan H-L, Haffner MC, Esopi DM, Vaghasia AM, Giannico GA, Ross HM, et al. Prostate adenocarcinomas aberrantly expressing p63 are molecularly distinct from usual-type prostatic adenocarcinomas. Mod Pathol [Internet]. 2015;28:446–56. Available from: http://dx.doi.org/10.1038/modpathol.2014.115

35. Jiang Z, Li C, Fischer A, Dresser K, Woda BA. Using an AMACR (P504S)/34betaE12/p63 cocktail for the detection of small focal prostate carcinoma in needle biopsy specimens. Am J Clin Pathol [Internet]. 2005;123:231–6. Available from: http://dx.doi.org/10.1309/1g1nk9dbgfnb792l

36. Furusato B, Tan S-H, Young D, Dobi A, Sun C, Mohamed AA, et al. ERG oncoprotein expression in prostate cancer: clonal progression of ERG-positive tumor cells and potential for ERG-based stratification. Prostate Cancer Prostatic Dis [Internet]. nature.com; 2010;13:228–37. Available from: http://dx.doi.org/10.1038/pcan.2010.23

37. Chaux A, Albadine R, Toubaji A, Hicks J, Meeker A, Platz EA, et al. Immunohistochemistry for ERG expression as a surrogate for TMPRSS2-ERG fusion detection in prostatic adenocarcinomas. Am J Surg Pathol [Internet]. ncbi.nlm.nih.gov; 2011;35:1014–20. Available from: http://dx.doi.org/10.1097/PAS.0b013e31821e8761

38. Tomlins SA, Alshalalfa M, Davicioni E, Erho N, Yousefi K, Zhao S, et al. Characterization of 1577 primary prostate cancers reveals novel biological and clinicopathologic insights into molecular subtypes. Eur Urol [Internet]. Elsevier; 2015;68:555–67. Available from: http://dx.doi.org/10.1016/j.eururo.2015.04.033

39. Baydar DE, Kulac I, Gurel B, De Marzo A. A case of prostatic adenocarcinoma with aberrant p63 expression: presentation with detailed immunohistochemical study and FISH analysis. Int J Surg Pathol [Internet]. 2011;19:131–6. Available from: http://dx.doi.org/10.1177/1066896910379478

40. Giannico GA, Ross HM, Lotan T, Epstein JI. Aberrant expression of p63 in adenocarcinoma of the prostate: a radical prostatectomy study. Am J Surg Pathol [Internet]. 2013;37:1401–6. Available from: http://dx.doi.org/10.1097/PAS.0b013e31828d5c32

41. Torres A, Alshalalfa M, Davicioni E, Gupta A, Yegnasubramanian S, Wheelan SJ, et al. ETS2 is a prostate basal cell marker and is highly expressed in prostate cancers aberrantly expressing p63. Prostate [Internet]. 2018; Available from: http://dx.doi.org/10.1002/pros.23646

42. De Marzo AM, Marchi VL, Epstein JI, Nelson WG. Proliferative inflammatory atrophy of the prostate: implications for prostatic carcinogenesis. Am J Pathol [Internet]. 1999;155:1985–92. Available from: http://dx.doi.org/10.1016/S0002-9440(10)65517-4

43. Zha S, Yegnasubramanian V, Nelson WG, Isaacs WB, De Marzo AM. Cyclooxygenases in cancer: progress and perspective. Cancer Lett [Internet]. Elsevier; 2004;215:1–20. Available from: http://dx.doi.org/10.1016/j.canlet.2004.06.014

44. Robinson D, Van Allen EM, Wu Y-M, Schultz N, Lonigro RJ, Mosquera J-M, et al. Integrative Clinical Genomics of Advanced Prostate Cancer. Cell [Internet]. 2015;162:454. Available from: http://dx.doi.org/10.1016/j.cell.2015.06.053

45. Cancer Genome Atlas Research Network. The Molecular Taxonomy of Primary Prostate Cancer. Cell [Internet]. 2015;163:1011–25. Available from: http://dx.doi.org/10.10167j.cell.2015.10.025

46. Armenia J, Wankowicz SAM, Liu D, Gao J, Kundra R, Reznik E, et al. The long tail of oncogenic drivers in prostate cancer. Nat Genet [Internet]. Nature Publishing Group; 2018 [cited 2018 Apr 3];1. Available from: https://www.nature.com/articles/s41588-018-0078-z

47. Quigley DA, Dang HX, Zhao SG, Lloyd P, Aggarwal R, Alumkal JJ, et al. Genomic Hallmarks and Structural Variation in Metastatic Prostate Cancer. Cell [Internet]. 2018;174:758–69.e9. Available from: http://dx.doi.org/10.1016/j.cell.2018.06.039

48. Wu Y-M, Cieślik M, Lonigro RJ, Vats P, Reimers MA, Cao X, et al. Inactivation of CDK12 Delineates a Distinct Immunogenic Class of Advanced Prostate Cancer. Cell [Internet]. 2018;173:1770–82.e14. Available from: http://dx.doi.org/10.1016/j.cell.2018.04.034

49. Viswanathan SR, Ha G, Hoff AM, Wala JA, Carrot-Zhang J, Whelan CW, et al. Structural Alterations Driving Castration-Resistant Prostate Cancer Revealed by Linked-Read Genome Sequencing. Cell [Internet]. 2018; Available from: http://dx.doi.org/10.1016/j.cell.2018.05.036

50. Rebbeck TR. Prostate Cancer Disparities by Race and Ethnicity: From Nucleotide to Neighborhood. Cold Spring Harb Perspect Med [Internet]. 2018;8. Available from: http://dx.doi.org/10.1101/cshperspect.a030387

51. Wallace TA, Prueitt RL, Yi M, Howe TM, Gillespie JW, Yfantis HG, et al. Tumor immunobiological differences in prostate cancer between African-American and European-American men. Cancer Res [Internet]. 2008;68:927–36. Available from: http://dx.doi.org/10.1158/0008-5472.CAN-07-2608

52. Reams RR, Renee Reams R, Agrawal D, Davis MB, Yoder S, Odedina FT, et al. Microarray comparison of prostate tumor gene expression in African-American and Caucasian American males: a pilot project study [Internet]. Infectious Agents and Cancer. 2009. Available from: http://dx.doi.org/10.1186/1750-9378-4-s1-s3

53. Powell IJ, Dyson G, Land S, Ruterbusch J, Bock CH, Lenk S, et al. Genes associated with prostate cancer are differentially expressed in African American and European American men. Cancer Epidemiol Biomarkers Prev [Internet]. 2013;22:891–7. Available from: http://dx.doi.org/10.1158/1055-9965.EPI-12-1238

54. Maynard JP, Ertunc O, Kulac I, Baena-Del Valle JA, De Marzo AM, Sfanos KS. IL8 Expression Is Associated with Prostate Cancer Aggressiveness and Androgen Receptor Loss in Primary and Metastatic Prostate Cancer. Mol Cancer Res [Internet]. 2020;18:153–65. Available from: http://dx.doi.org/10.1158/1541-7786.MCR-19-0595

55. De Marzo AM, Platz EA, Sutcliffe S, Xu J, Grönberg H, Drake CG, et al. Inflammation in prostate carcinogenesis. Nat Rev Cancer [Internet]. 2007;7:256–69. Available from: http://dx.doi.org/10.1038/nrc2090

56. Sfanos KS, Yegnasubramanian S, Nelson WG, De Marzo AM. The inflammatory microenvironment and microbiome in prostate cancer development. Nat Rev Urol [Internet]. 2017; Available from: http://dx.doi.org/10.1038/nrurol.2017.167

57. Liu X, Grogan TR, Hieronymus H, Hashimoto T, Mottahedeh J, Cheng D, et al. Low CD38 Identifies Progenitor-like Inflammation-Associated Luminal Cells that Can Initiate Human Prostate Cancer and Predict Poor Outcome. Cell Rep [Internet]. 2016;17:2596–606. Available from: http://dx.doi.org/10.1016/j.celrep.2016.11.010

58. Iwao-Koizumi K, Matoba R, Ueno N, Kim SJ, Ando A, Miyoshi Y, et al. Prediction of docetaxel response in human breast cancer by gene expression profiling. J Clin Oncol [Internet]. 2005;23:422–31. Available from: http://dx.doi.org/10.1200/JC0.2005.09.078

59. Arai T, Miyoshi Y, Kim SJ, Akazawa K, Maruyama N, Taguchi T, et al. Association of GSTP1 expression with resistance to docetaxel and paclitaxel in human breast cancers. Eur J Surg Oncol [Internet]. 2008;34:734–8. Available from: http://dx.doi.org/10.1016/j.ejso.2007.07.008

60. Miyake T, Nakayama T, Naoi Y, Yamamoto N, Otani Y, Kim SJ, et al. GSTP1 expression predicts poor pathological complete response to neoadjuvant chemotherapy in ER-negative breast cancer [Internet]. Cancer Science. 2012. page 913–20. Available from: http://dx.doi.org/10.1111/j.1349-7006.2012.02231.x

61. Eralp Y, Keskin S, Akisik E, Akisik E, Igci A. Predictive role of midtreatment changes in survivin, GSTP1, and topoisomerase 2α expressions for pathologic complete response to neoadjuvant …. American journal of [Internet]. journals.lww.com; 2013; Available from: https://journals.lww.com/amjclinicaloncology/fulltext/2013/06000/Predictive_Role_of_Midtreatment_Changes_in.1.aspx

62. Işeri OD, Kars MD, Gündüz U. Two different docetaxel resistant MCF-7 sublines exhibited different gene expression pattern. Mol Biol Rep [Internet]. 2012;39:3505–16. Available from: http://dx.doi.org/10.1007/s11033-011-1123-5

